# Risk Factors and Pivotal Periods: Marital Status over 28 Years for Parents of Individuals with Autism

**DOI:** 10.1101/2022.02.28.22271595

**Authors:** Niki Bahri, Kyle Sterrett, Catherine Lord

**Author notes:** **Address correspondence to:** Catherine Lord, Semel Institute for Neuroscience and Human Behavior, University of California, Los Angeles, 760 Westwood Plaza 68-217, Los Angeles, CA 90024. **Data Sharing Statement:** De-identified data will be made available upon request. **Contributors’ Statement:** Ms. Bahri conceptualized and designed the study, interpreted the data, prepared the first draft of the manuscript and reviewed and revised the manuscript. Dr. Lord conceptualized and designed the study, interpreted the data and critically reviewed the manuscript for important intellectual content. Dr. Sterrett carried out the formal analyses and interpretation of the data, contributed to the first draft of the manuscript, and reviewed and revised the manuscript. All authors approved the final manuscript as submitted and agree to be accountable for all aspects of the work.

## Abstract

Longitudinal, prospective analyses of marital status in parents of individuals with autism are needed. We describe the timing of divorce, and the factors that contribute to divorce in a longitudinal sample of families of individuals with autism. Participants included parents of 219 children, initially referred for autism and other developmental delays, followed to age 30 years. Approximately 36% of individuals with autism in our sample experienced a parental divorce by age 30. Higher rates of divorce were associated with maternal education, race and age at child’s birth, as well as autism symptom severity and diagnosis. Divorces were most common in early years (under age 5) and also in the teenage years and beyond (over age 15). After age 15, higher risk was associated with higher cognitive ability and daily living skills, and being a multiplex family. Results suggest that divorce risk in families of children with autism remains high through childhood into early adulthood. Understanding factors related to changes in marital status may help us better support families across time.

---

Raising a child with autism can be a source of considerable stress for caregivers (Hayes et al., 2013). Unique parental related stressors such as children’s behavior problems, changes in family routines to accommodate children’s needs, struggling to navigate the service system, and constraints on parent’s time, affect caregivers’ mental health and their relationships (Iadarola et al., 2019; Schieve et al., 2007). Caregivers of children with autism report reduced quality of life, and high stress, depression, and anxiety due to the ongoing demands of care (Brobst et al., 2009; Brown et al., 2020; Higgins et al., 2005; Schnabel et al., 2020). They also report lower marital and relationship satisfaction when compared to parents of typically developing children (Brobst et al, 2009; Higgins et al., 2005). Further, higher levels of mental health related concerns and lower relationship satisfaction have been negatively associated with divorce rates in these families (Risdal & Singer, 2004; Seltzer et al., 2011).

## Impact of Divorce

The disruption to the family caused by divorce, or parental separation, can impact children’s development and academic progress. Across multiple samples, children whose parents separated saw increased school related difficulties; this is especially the case when some academic difficulties are already present (Babalis et al., 2014; Bernardi et al., 2014; Nusinovici et al., 2018). Though, the impact of divorce appears to be temporally associated with the separation and becomes less apparent as children age (Clare-Stewart et al., 2000). This indicates that the transition, and associated disruptions, are a major contributor to the negative effects seen in children experiencing a parental divorce. However, the effects of divorce in childhood do have the potential to carry over into adulthood, even affecting physical health outcomes (Thomas & Högnas, 2015). Thus, it is important to empirically evaluate whether families of children with autism are at higher risk for experiencing divorce and its potentially negative developmental consequences.

## Estimates of Divorce Rates

Comparative samples including data from both typically developing children and children with disabilities are limited. Overall, when comparing the divorce rates of parents of children with disabilities to the general population of parents, the findings have been inconclusive. Some studies have found higher rates of marital instability in families of children with disabilities compared to families without a child with a disability (Freeman et al., 2010; Hatton et al., 2010; Reichman et al., 2004). Others have found comparable divorce rates between the two groups (Namkung et al., 2015; Seltzer et al., 2001).

The data on divorce rates in families of children with autism has been similarly inconsistent. Early media reports claimed, without empirical support, that up to 80% of parents raising a child with autism would divorce (Freedman et al., 2012). Though almost all empirically derived rates have been much lower; most hover around 30% (Baeza-Velasco et al., 2013; Harley et al., 2010; Hoover & Kaufman, 2018).

When comparing the divorce rates of families of children with autism with the general population cross-sectionally, some studies found slightly higher rates in the autism groups. Other studies have found comparable rates across the autism and comparison groups. For example, analyses from a national survey indicated that children with autism are at a moderately increased risk of parental divorce compared to their peers (28% v. 20%) (Hoover & Kaufman, 2018). However, in a sample of 77,911 parents with children 3-17 years old (913 with autism diagnoses), Freedman et al. (2012) found no evidence that children with autism were at increased risk of experiencing parental separation.

Given these conflicting findings, and the fact that the impact of autism on individuals, parents, and families changes over time, longitudinal studies can provide more nuanced information that builds on cross-sectional “snapshots” (Namkung et al., 2015). Despite the importance of longitudinal data to thoroughly investigate changes, there are few well-characterized ADS longitudinal samples that have reported marital status data. Baeza-Velasco et al. (2013) did not include a comparison group, but reported a divorce rate of 25.3% over a 10-year period beginning when children were approximately 5 years old (Baeza-Velasco et al., 2013). In another sample, the prevalence of divorce was significantly higher among the parents of children with autism than parents of a well-matched sample of children without a disability (23.5% vs. 13.8%; Hartley et al., 2010). Though Hartley et al. (2010) used a longitudinal sample, the wide age range (14-56 years) of individuals, and the lack of repeated in-depth, behavioral data limits the ability to assess the role of age interacting with other factors, including parent and participant characteristics. These two studies are to our knowledge the only longitudinal investigations of changes in marital status in caregivers of children with autism.

## The Impact of Parent, Family and Individual Factors

Family systems are complex. It is important to better understand how familial, parent and child level factors interact with each other within those systems, and how they influence the likelihood of experiencing a divorce over and beyond the child’s disability status alone. Overall, the relationship between divorce and familial, child and parental factors in longitudinal samples of individuals with autism have been inconsistent. Hartley et al. (2010) found that divorce was more frequent when children with autism were born later in the birth order, and when mothers were younger. Baeza-Velasco et al. (2013) found no significant differences between parents who remained together and parents who separated based on any sociodemographic variables. In both studies, clinical characteristics (e.g., severity of early autism symptoms) were unrelated to divorce (Baeza-Velasco et al., 2013; Hartley et al., 2010). Continued evaluation of parent, family and individual factors that may contribute to divorce risk remains important.

## Pivotal Periods in Risk for Divorce

Beyond evaluating the influence of family, parent and individual factors on divorce rates, longitudinal data also provide the opportunity to identify vulnerable periods in marriages. Previous divorce data among the general population show that parents are at greatest risk of divorce before their children are teenagers (Bramlett & Mosher, 2002; Hirschberger et al., 2009). While parenting demands may decrease for caregivers in the general population as their children get older, many children with autism require substantial support into adulthood (Burke & Heller, 2016). Two studies have evaluated the timing of divorce in parents of children with autism, and found that risk for divorce remains high into early adulthood (Baeza-Velasco et al., 2013; Hartley et al., 2010). This supports the idea that continued caregiving demands in aging parents may contribute to higher divorce rates in families of adolescents and adults with autism. Parents of children with autism are also likely to divorce earlier in their child’s development than parents of children without a disability (Hartley et al., 2010). Again, suggesting that the timing of parental divorce could differ for families of children with autism, with vulnerable periods occurring earlier, as well as later, in the life course. Identifying vulnerable periods for divorce provides the opportunity to support families when it is most needed (Bramlett and Mosher, 2002). For providers, a realistic recognition of the relationship between autism and marital status, as well as the influence of other clinical and sociodemographic characteristics on divorce and its timing, is important to improving the quality of care provided to families.

The studies described above provide important information regarding parent marital status within relatively brief time periods. Here, we evaluate changes in marital status over a 28-year period (from child age 2 to 30) among parents of a well-described, age-homogeneous, longitudinal sample of 253 individuals with autism and other early-diagnosed developmental disabilities. We consider both the attributes of the individuals with autism and parent and family factors associated with divorce, and investigate whether there are pivotal time periods in marital status change.

## Methods

### Participants

Participants were drawn from the Autism Early Diagnosis Cohort (see Lord et al., 2006; Anderson et al., 2007); a sample of consecutive community-referrals, all under age 3, who were referred for suspected autism from two programs (North Carolina and Chicago). At the initial visit, 192 of the children received an autism diagnosis and 21 children had developmental delay, but did not meet autism diagnostic criteria. Forty additional participants with previous autism diagnoses in early childhood joined at approximately age 10 (Michigan) (see Anderson et al., 2014).

From the full sample of 253 families, 34 children living with an unmarried (n=27), widowed (n=1) or parent with unknown marital status (n=6) were excluded, resulting in a final sample of 219 participants. The sample includes 9 foster or adoptive parents. In the active participants, who are currently enrolled in the study, there have been no reported parental divorces from age 25 to 30, with 2 parental deaths and 3 remarriages. Data were collected from 1992 through 2020 and parents and caregivers reported marital status approximately every 6 months during that time (see Figure 1).

**Figure 1.**
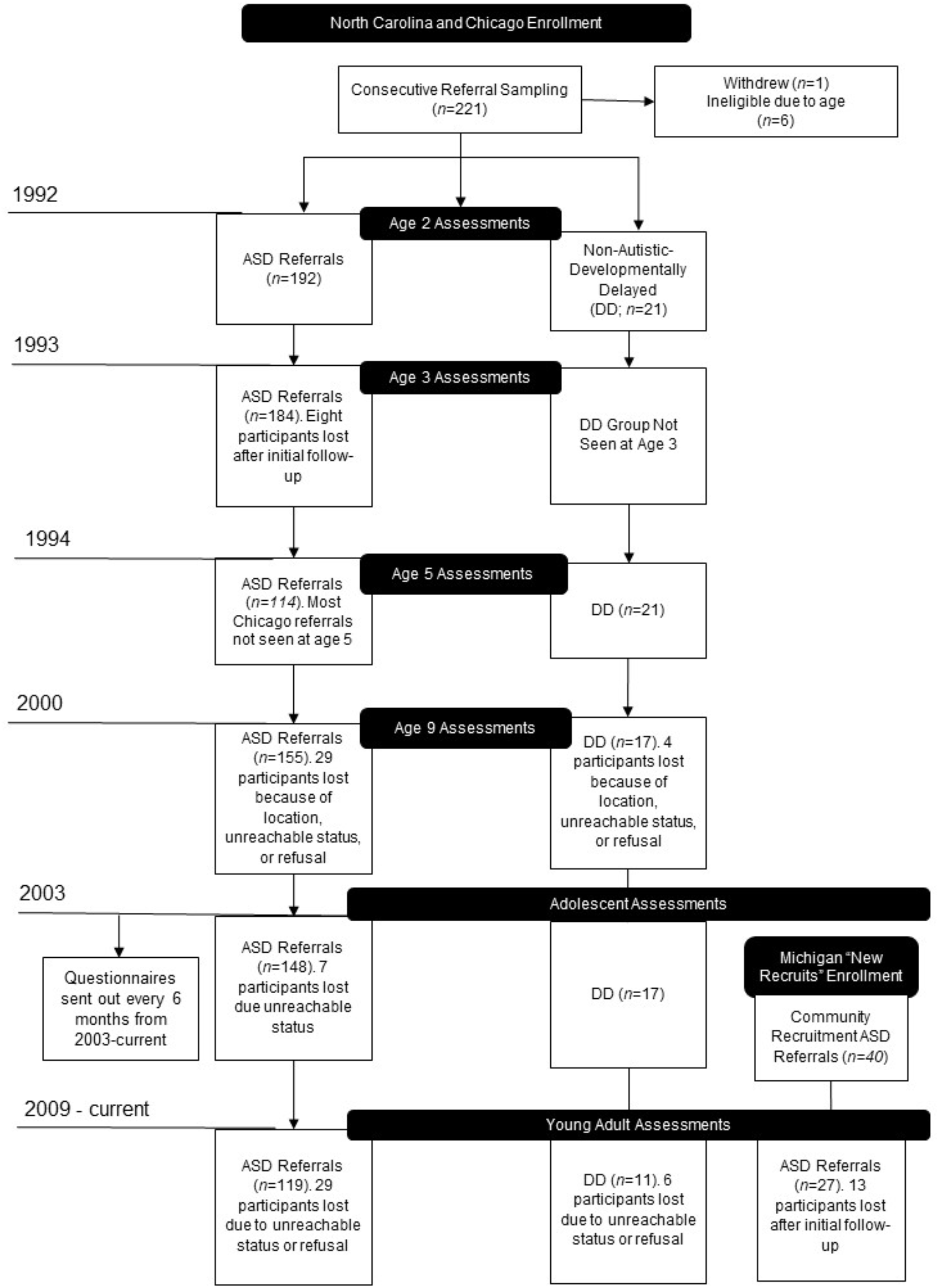
Consort Diagram for Study Flow.

### Procedures

Families completed in-person assessments at approximately ages 2, 3 (autism referrals), 5 (North Carolina only), 9, 14, 18, 21, and 25. All behavioral measures were collected at in-person assessments. At these in-person visits, best-estimate formal diagnoses were made by experienced clinicians, blinded to prior diagnoses, using standardized instruments and clinical judgment. Clinicians used all available information, including standardized diagnostic instruments, cognitive testing, and parent reports of adaptive behavior to make the diagnoses at each visit. Across the 8 visits, 48 participants never received an autism spectrum disorder (ASD) diagnosis (referred to later as “Never ASD”) and 171 received an ASD diagnosis during at least one assessment visit (referred to later as “Ever ASD”).

In addition to the in-person assessments where behavioral measures were administered, participants completed self- and caregiver-report psychological measures approximately every 6 months. Informed consent was obtained from all participating families and, whenever possible, individuals themselves. The research was approved by each site’s Institutional Review Board.

### Instruments

#### Marital Status

Marital status information was gathered through interviews, and self-reported demographic forms, at every data point. Parents were asked to report marital status (single, widowed, remarried, divorced or separated) and the date of separation, divorce, or remarriage when applicable. When reported, the date of separation was used to signify the end of a couple’s relationship, as has been done in previous studies (e.g., Baeza-Velasco et al., 2013).

#### Characteristics of Families

Familial and child characteristics were collected through the same procedures described above. Familial characteristics included: mother’s race (white, non-Hispanic compared to people of color), education (less than a four-year college degree compared to four-year college or graduate degree) and age at child’s birth, number of children in the family, having another child with autism, and original study site (North Carolina, Chicago, Michigan). We also examined the child’s gender and birth order (only, youngest, middle or oldest child).

#### Behavioral Measures

Autism symptom severity was measured by the Autism Diagnostic Observation Schedule (ADOS), a standardized observation of social and communicative behavior with good reliability and validity (Lord et al., 2012). Calibrated Severity Score (CSS), including CSS Social Affect (CSS SA), and CSS Restricted and Repetitive Behaviors (CSS RRB) scores were used rather than raw scores, in line with previous recommendations (Gotham et al., 2009). Verbal intelligence quotients (VIQ) and non-verbal intelligence quotients (NVIQ) were gathered from the Wechsler Abbreviated Scale of Intelligence, the Differential Ability Scales, and the Mullen Scales of Early Learning following a standard hierarchy based on participants age and ability (Anderson et al., 2014; Elliot, 2007; McCrimmon et al., 2013; Mullen, 1995). Daily living skills scores were collected using the Vineland Adaptive Behavior Scales, a parent interview that primarily assesses three domains: Communication, Daily Living, and Socialization (Sparrow et al., 2005).

As behavioral scores in this sample have been shown to be relatively stable across time, (see Gotham et al., 2012) in order to maximize the amount of available data, age 9 scores were used for ADOS and Vineland scores; when data were missing at age 9, age 5 or the next available assessment was used. For cognitive scores, individual’s highest VIQ and NVIQ scores, across any timepoint, were used.

### Data analysis plan

Demographic variables were compared across marital status (married or divorced) using t-tests for continuous variables and chi-square tests for categorical variables. The primary analyses were carried out using the Survival package in R, version 4.0.2 (R Core Team, 2020; Therneau & Grambsch, 2000). Kaplan-Meyer survival curves were fit to evaluate the overall risk of divorce in the sample. Cox proportional-hazards models were then fit separately for child (gender, birth order, ADOS CSS SA and CSS RRB, ever ASD diagnosis, highest VIQ and daily living scores) and family factors (mother’s age, education, race and number of children) to determine which variables to include in the final model. Non-significant variables from the separate child and parent models were dropped in the final, combined model. In the separate child and parent models, variables were selected based on their substantive importance and relevance to the dependent variable and their inclusion in prior literature on marital status in autism samples (Baeza-Velasco et al., 2013; Hartley et al., 2010). Sample sizes varied slightly based on missing data patterns. Last, a piecewise exponential survival analysis was conducted, using a generalized linear model to compare whether the odds varied across different age ranges: 0 to 5 years of age, 5 to 10 years, 10 to 15 years and older than 15 years. Age 5 to 10 years old was used as the reference group. Race, education and diagnosis controlled for in this model. In order to compare demographic and child characteristics across the four age bins, chi-square tests and ANOVA were used. Significant ANOVAs were followed up with pairwise contrasts between the groups with a Tukey correction applied.

### Community Involvement

There was no community involvement in the reported study.

## Results

### Attrition

Of the 219 included families, 108 had censored data on marital status by age 30, either because of non-response or attrition. Independent samples t-tests for continuous data and chi-square tests for categorical data were used to determine whether there were systematic differences in those with and without complete data. Complete marital status data was associated with higher parental education (p=.04) and having fewer children in the family (p=.04), but not race, child gender, site, CSS scores, mother’s age, father’s age, NVIQ, VIQ or Ever ASD diagnosis (all p>.05). Attrition in the full sample has been associated with both lower parent education and African-American race (see Pickles et al., 2020).

### Early Marital Status

Being unmarried when their children were 2 years old was associated with lower maternal age (p<.001), lower parental education (p<.001), race (p <.001) and North Carolina study site (p<.001), but no child characteristics. See Table 1.

**Table 1.** Sample Descriptive Information

|  | **All** | **Divorced** | **Married** |
| --- | --- | --- | --- |
| *n* | 219 | 67 | 152 |
| **Mother** |  |  |  |
| Age in years at proband birth (M, SD) | 29.90 (5.73) | 28.00 (5.27) | 30.7 (5.75) |
| Range | 17-57 | 18-36 | 17-57 |
| White, non-Hispanic (%) | 76.6 | 64.2 | 82.1 |
| < High school education (%) | 2.1 | 3.4 | 1.5 |
| High school graduate (%) | 16.3 | 15.5 | 16.7 |
| Some college/Bachelor's degree (%) | 27.9 | 43.1 | 21.2 |
| Four year college degree/Post<br>Bachelor's/Graduate degree (%) | 53.5 | 37.9 | 60.6 |
| **Father** |  |  |  |
| Age in years at proband birth (M, SD) | 31.84 (5.51) | 30.3 (6.03) | 32.5 (5.19) |
| Range | 14- 45 | 18-44 | 14-45 |
| **Child** |  |  |  |
| Male (%) | 79.9 | 82.1 | 78.9 |
| Birth Order<br>current |  |  |  |
| Only child (%) | 27.6 | 27.7 | 27.6 |
| First born (%) | 38.7 | 40.0 | 38.2 |
| Middle child (%) | 17.05 | 7.7 | 21.1 |
| Youngest child (%) | 16.6 | 24.6 | 13.2 |
| Sibling with autism current (%) | 12.8 | 13.4 | 12.5 |
| Total # of children, current (M, SD) | 1.80 (1.31) | 1.92 (1.37) | 1.74 (1.29) |
| Range | 0-7 | 0-5 | 0-7 |
| Verbal IQ age 9 (M, SD) | 59.46 (37.64) | 57.0 (35.3) | 60.4 (38.7) |
| Non-verbal IQ age 9 (M, SD) | 66.81 (33.11) | 65.2 (32.6) | 67.2 (33.4) |
| ADOS Calibrated Severity Scores age<br>9 (M, SD) | 6.06 (2.92) | 6.57 (2.74) | 5.84 (2.98) |
| Vineland Daily Living Skills Standard<br>Score age 9 (M, SD) | 63.27 (16.60) | 60.8 (15.1) | 64.5 (17.2) |

*Note.* Behavioral data was used from the age 9 visit or the next closest visit if not available. Variables under “divorced” category are based on raw data up until participant age 30. The Married category includes participants whose parents were married until age 30 or their final available data point. IQ= Intelligence Quotient, M= Mean, SD= Standard Deviation.

### Overall Rates of Divorce

See Figure 2 for survival curve graphs across all variables of interest; in these graphs, survival refers to not having experienced a divorce. A graph of the overall risk can be found in Appendix A. Estimated risk of experiencing a parental divorce was approximately 36% by age 30.

**Figure 2.**
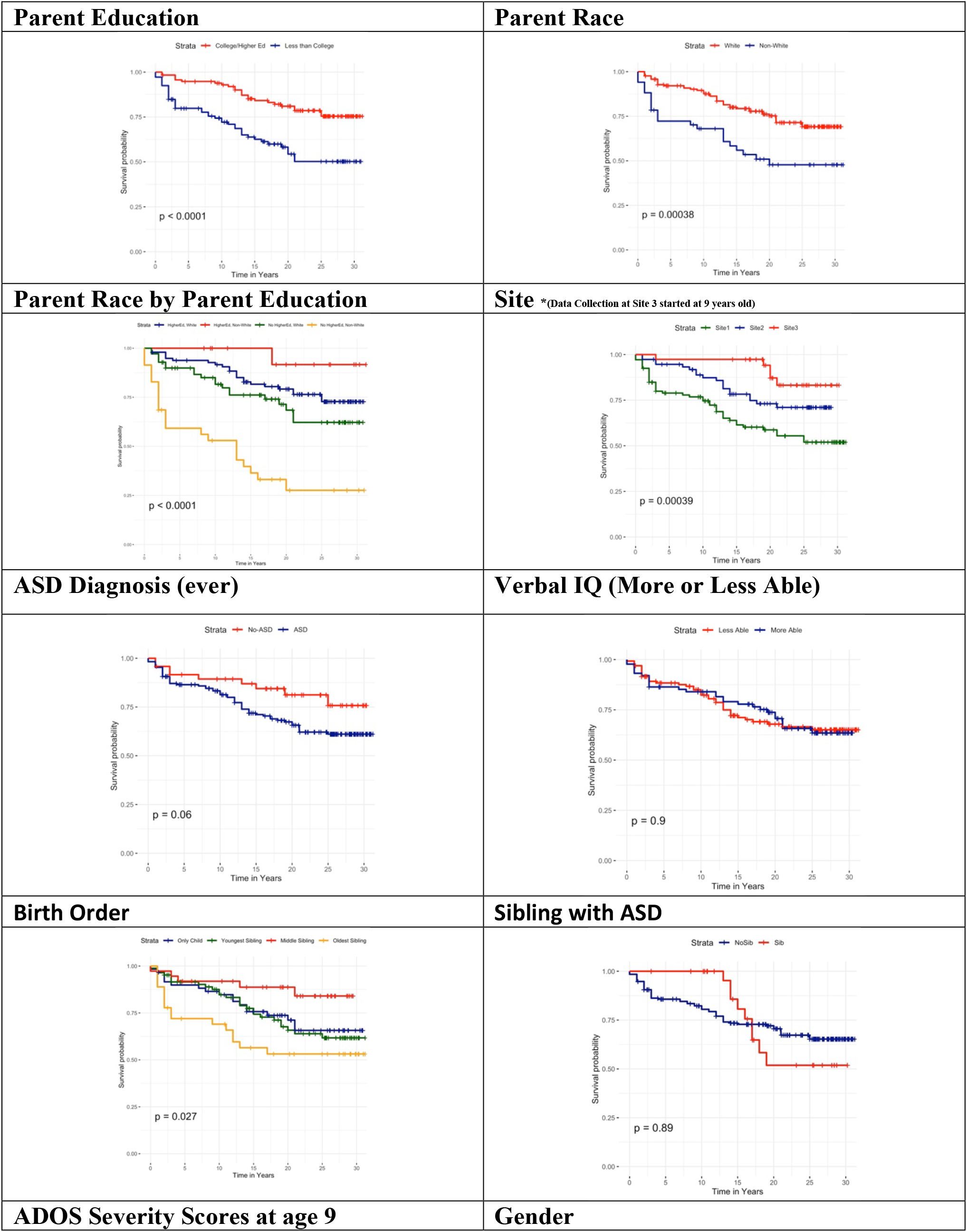
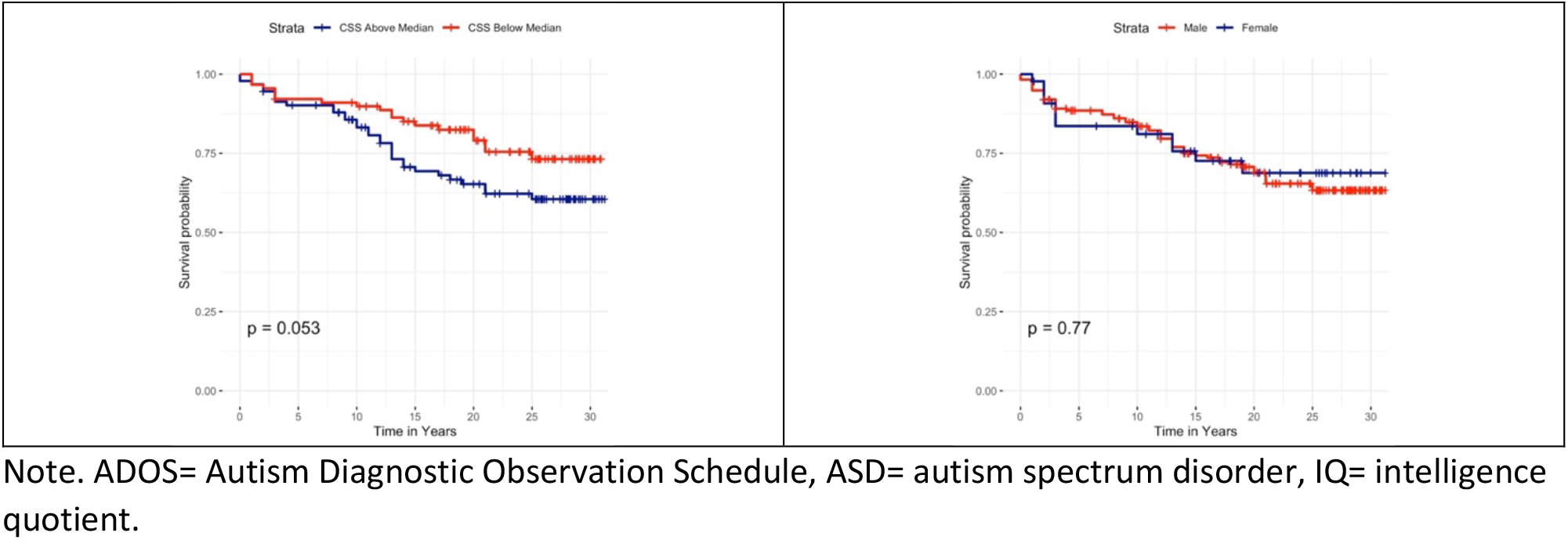
Survival Curves Split by Clinical and Socio-Demographic Factors.

### Child Characteristics Univariate Models

In the univariate models, there was a small difference in the marriage survival rates for families with “Ever ASD” and “Never ASD” participants. The “Ever ASD” group were 15% more likely to experience a divorce by age 30 (log-rank; p= .059), as were those with higher autism severity (CSS of 7 or greater), with a 13% higher risk than those with CSS less than 7 (log rank; p=.053). Families with only children were 13%, 15% and 15% more likely to experience a divorce than youngest, middle and oldest children in the birth order, respectively (log-rank; p=.02). Divorce rates were not different based on participants’ cognitive ability (log-rank; p=.92) nor participants’ gender (log rank; p=.73).

Having a sibling with autism (multiplex families) violated the non-proportional hazards assumption. There was a sharp increase in risk for divorce around 13 years old in those with a sibling with autism, and the risk in multiplex families surpassed risk in families with only one child with autism at around this point. This finding is visualized in Figure 3 but not tested using a log-rank test.

### Parent Characteristics Univariate Models

In the univariate models, approximately 49% of parents with less than a college education compared to 26% of parents with a college education experienced a divorce (log-rank; p<.0001). Further, 31% of white mothers compared to 52% of mothers of color experienced a divorce (log-rank; p<.0001). Those at highest risk for divorce were mothers of color with less than a college education, with an estimated risk for divorce of 71% by the end of the study period (log-rank; p<.0001).

### Preliminary Multivariate Models

In the preliminary child variable model, there was a significant effect of birth order; only children were at highest risk for experiencing a divorce when compared to youngest (b=-0.87, se=.42, p=.007), middle (b=-1.25, se=.29, p= .003) and oldest children (b=-0.97, se=.38, p= .02). All other coefficients were non-significant. However, there was some signal that an autism diagnosis leads to increased risk of experiencing a divorce (b=0.82, se=.45, p=.06).

In the preliminary parent variable model, risk of divorce was significantly higher in parents with a high school education or less (b=0.86, se=.29, p-.003), in mothers of color (b=.75, se=.28, p=.006) and in younger mother’s (b=-.05, se=.02, p=.02). The effect of number of children in the family was non-significant (p=.23). We also tested the interaction between race and education status, which was significant (b=2.35, se= 1.08, p=.03). The effect of race on risk for divorce is stronger in parents with less education. Parameters for the Child and Parent models are included in Appendix B.

### Parent and Child Characteristics Combined Model

The final combined model included autism diagnosis, birth order, age, site and the interaction between mother’s race and education. There was a site effect with those in Michigan being at reduced risk for divorce compared to Chicago (b=-.64, se=.32, p=.05), and non-significantly reduced risk compared to North Carolina (b=-1.45, se=.77, p=.06). This is likely because Michigan families entered the study in late childhood, after one of the periods of greatest changes in marital status. There was a significant effect of mother’s age (b=-.05, s=.02, p=.02) in the same direction as described above where younger mothers were at higher risk for divorce. The significant interaction between mother’s race and education remained significant in this model as well (b=2.21, se=1.08, p=.04). The only difference that remained in the birth order variable was an increased risk for experiencing a divorce in only children compared to middle children (b=-.87, se= 41, p=.03). Ever ASD diagnosis was no longer significant (p>.05). See Table 2 for full model parameters.

**Table 2.** Cox Regression- Combined Model

| **Variable** | **Hazard Ratio** | **95% CI** | **P-value** |
| --- | --- | --- | --- |
| Race | 0.27 | [0.03, 2.03] | 0.20 |
| Mother's Education | 1.41 | [0.70, 2.83] | 0.02 |
| Race*Education | 9.08 | [1.09, 75.38] | 0.04 |
| Mother's Age | 0.95 | [0.90, 0.99] | 0.01 |
| ASD Ever |  |  |  |
| Birth Order |  |  |  |
| Youngest Child | 0.71 | [0.36, 1.41] | 0.31 |
| Middle Child | 0.42 | [0.18, 0.94] | 0.06 |
| Oldest Child | 0.55 | [0.23, 1.29] | 0.36 |
| Site |  |  |  |
| Chicago | 0.53 | [0.28, 0.99] | 0.05 |
| North Carolina | 0.23 | [0.05, 1.05] | 0.06 |

*Note.* Reference group for Race is white and the comparison group is people of color, reference group for Parent Education is less than college education and the comparison group is college education or higher, reference group for ASD Ever is No ASD Diagnosis, reference group for Birth Order is Only Child, and reference group for Site is Michigan.

### Pivotal Periods in Divorce

The proportion of families who experienced a divorce during each of the time periods were: 12.5% from 0-5 years, 4.7% from 5-10 years, 9.4% from 10 to 15 years and 8.6% from 15 to 30. The period between 5 and 10 years old had lower parental divorce risk compared to both 0 to 5 years old (OR=2.25; p=.04) and the older than 15 years old group (OR=2.30; p=.05). There was also a non-significant difference between the 5 to 10 years and 10 to 15 years groups in the same direction (OR=2.17; p=.06). See Appendix A and Appendix C for more information.

#### Factors Associated with Pivotal Periods in Divorce

When comparing the characteristics of individuals who experienced a divorce across the age bins, the ANOVA for NVIQ was significant (F (3, 60) = 3.36, p=.02); differences in NVIQ were seen between families who divorced between 15-30 years and 10-15 years (marginal mean difference= 29.99, 95% CI [-0.08, 60.07]; p=.05) as well as between 15-30 years and 5-10 years (marginal mean difference= 37.68, 95% CI [2.28, 73.08]; p= .03). Individuals who were teens and young adults when their parents divorced had the highest NVIQ scores.

The ANOVA for daily living skills was also significant (F (3, 59) = 3.45, p=.02); pairwise comparisons showed differences between those participants who experienced divorce between 15-30 years old and 10-15 years old (marginal mean difference= 14.29, 95% CI [.06, 28.51]; p=.05). Again, those in the 15-30-year-old group had the highest daily living scores. All other pairwise comparisons and ANOVAs were non-significant.

While we are limited by a small sample, there was a signal for a difference between having a sibling with autism or not (multiplex versus simplex families) on divorce risk across age bins (χ^2^ (3, 67) = 7.25, p=.06). There was an unexpected crossover effect at about age 13 (see Fig. 2). No parents of individuals with two or more siblings with autism divorced between 0-10 years, whereas 46% of divorces in families with non-autistic siblings occurred during the 0-10 years period. 33% of families with two or more siblings with autism divorced after age 10. Sibling status itself did not appear to be driving differences in participants characteristics; there were no differences between child-participants with siblings with autism and those without siblings with autism on ADOS CSS, daily living, NVIQ or VIQ scores.

## Discussion

The risk of participants experiencing a divorce by 30 years old was 36%. This is consistent, though slightly higher, than the parental divorce rates reported in other longitudinal samples of children with autism, both approximately 25% (Baeza-Velasco et al., 2013; Hartley et al., 2010). There is a need to recruit more recent, well-characterized, samples to further validate and track these estimates over time.

One challenge, given the absence of a control group, is attempting to draw a direct comparison between the divorce rates in our sample and the prevalence of divorce in the general population, and those with disabilities other than autism. While many of the widely referenced divorce rates are dated, some recent studies have found estimates ranging from 22% in individuals with any developmental disability (Namkung et al., 2015) to 7.6% in a sample of individuals with Down Syndrome (Urbano & Hodapp, 2007). These differences may reflect temporal changes, reflecting that divorce rates have not been consistent in the last 10-20 years. They may also indicate that it is important to individually estimate the effects of various disability categories on divorce risk, rather than aggregating across disability categories.

The projection in our sample, and the others listed above, are all lower than national estimates suggesting nearly 48 percent of American marriages end in divorce (Cherlin, 2005). The most consistent and widely reported source for divorce rates in the general population during the time period of this study (1990-2020) are based on census data. However, the methods used in the formulation of these data do not allow for a direct comparison with our sample. Data from these sources are typically presented based on the number of individuals experiencing a divorce per 1000 individuals surveyed. Despite not being directly comparable to our sample, it is helpful to note that overall divorce rates (number of divorced people per 1000 surveyed) has been steadily declining across all three states; Illinois, North Carolina and Michigan, from where children in our sample were recruited (Center for Disease Control and Prevention, 2021). So, it is less likely that the divorces in later adolescence and adulthood that we observed are explained by an increase in rates in the general population in recent years.

### Factors that Influence Divorce Risk

The often drastically different estimates for divorce risk across samples highlight the importance of considering the influence sampling. The characteristics of the sample can influence divorce estimates. Consistent with prior literature (Aughinbaugh et al., 2013; Bulanda & Brown, 2007; Rotz, 2016) parental and family factors affected the likelihood of divorce in our sample. Families at highest risk for divorce tended to have less than a college education with mothers younger at the time of birth. There were also racial differences in risk for divorce that were primarily driven by differences between white and families of color who had lower maternal education. However, this finding should be treated as exploratory given the effective sample size becomes smaller when comparing across the four categories (i.e., white and people of color, less and more educated).

Parents of only children with autism were at highest risk of a divorce in comparison to children with siblings; whether this is because families experiencing difficulties are less likely to have subsequent children or whether the presence of other children protects against divorce through positive mechanisms such as increased family cohesion or reduction of stress about future caretaking is not clear (Marciano et al., 2014; Taanila et al., 1999). This finding contrasts with Hartley et al (2012) who found an elevated rate of divorce in those born later in the birth order. Though, it should be noted that their sample was considerably older on average. Overall, these findings point to the continued importance of measuring and evaluating the influence of factors at all levels of the family system on risk for divorce.

### Pivotal Periods for Divorce Risk

Risk for divorce within families is not a static process. As children develop and family dynamics change, so does divorce risk. In our sample, divorce risk was highest during two time periods: participant age 0 to 5 years old and greater than 15 years old. In the older age bin, risk was highest specifically for teens with higher cognitive ability and daily living skills. Our results corroborate that divorce in families of children with autism remains high from childhood into early adulthood (Hartley et al., 2012). A particularly interesting finding is that more divorces occurred in families with adolescents and adults with higher cognitive ability and better daily living skills. This suggests that the transition age period may be especially taxing on families of adolescents and young adults with more skills. It may be that the stress caused by the transition planning process, and apprehension about how to support independence in their children, results in marital discord (Sosnowy et al., 2017). In our sample, around 60% percent of more cognitively able adults lived with their parents, and though 80% had some employment, only 40% had full-time paid employment (Lord et al., 2020). This furthers the idea that even in those individuals with more skills, concerns around independence, and the level of support necessary to achieve independence, are a potential source of marital discord. Consistent with Hartley et al (2012) the divorce rates in individuals who have left their parents’ home was not impactful in reducing risk for divorce. Divorce rates of those who recently left home were comparable to the overall estimate (both near 40%) and only 10% of divorces occurred within 2 years of an adult child leaving home in our sample.

An unexpected finding was that no individuals with siblings with autism experienced a divorce between 0-10 years, whereas 46% of divorces in those without a sibling with autism happened during the same time period. After age 13, we saw suggestions of a crossover effect, where divorces began to occur in those with siblings with autism. This finding, while it should be taken as preliminary, because of the limited sample size, could point to the resilience of multiplex families or the families need to maintain a stable household, at least until the children are older (Dissanayake et al., 2019).

### Limitations

First, one of the primary limitations of these data are that they are non-representative of the autism population as a whole. This sample is unique in many ways, and attempts to extrapolate these findings to the general population of individuals with autism should be done with caution. Further, because our sample was drawn from three regions across the US that varied substantially (e.g., some rural, urban and suburban areas), we felt that direct statistical comparisons of divorce rates from our sample to population data were not appropriate. However, we were able to compare individuals with autism to those with early developmental delays within the sample, where some evidence was found for increased divorce risk in families based on autism diagnosis. There was no effect of gender on risk for divorce, although due to the small number of females in the sample this warrants future attention. Further, attrition limited the studies power, and may have increased the number of false-negative results due to lack of sufficient power. Although the findings in this study are informative, it is crucial to keep in mind that divorce can be influenced by numerous circumstances that were un-measured in this study (Clarke & Berrington, 1998). Future research on parental marital status using other approaches and larger samples is much needed.

## Conclusion

Despite these limitations, these findings from a well-characterized sample of participants with autism and developmental disorders, and their families, from early childhood to early adulthood gives us insight into potential risk factors and sensitive time periods for parental divorce. It also provides information on the different factors that might affect marital status in early childhood versus when children approach adulthood.

This study further corroborates that we need to better support families with an individual with autism across the lifespan. Frontline health workers are often the most consistent and repeated source of such support for families of individuals with autism. Considering that mothers’ age and education were consistent divorce risk factors, increased focus on parent-mediated treatments that address family concerns as well child behaviors could be particularly important in providing support to young families that are traditionally underserved. Effective intervention and support, including those for transition age adolescents and young adults, should address characteristics related to autism and recognize the needs of parents as couples by providing resources, support and parent training if needed.

## Data Availability

All data produced in the present study are available upon reasonable request to the authors.

