## Supplemental Material for "Risk Factors and Pivotal Periods: Marital Status over 28 Years for Parents of Individuals with Autism"

**Appendix A**

Divorce Survival Curve

**Full Sample**

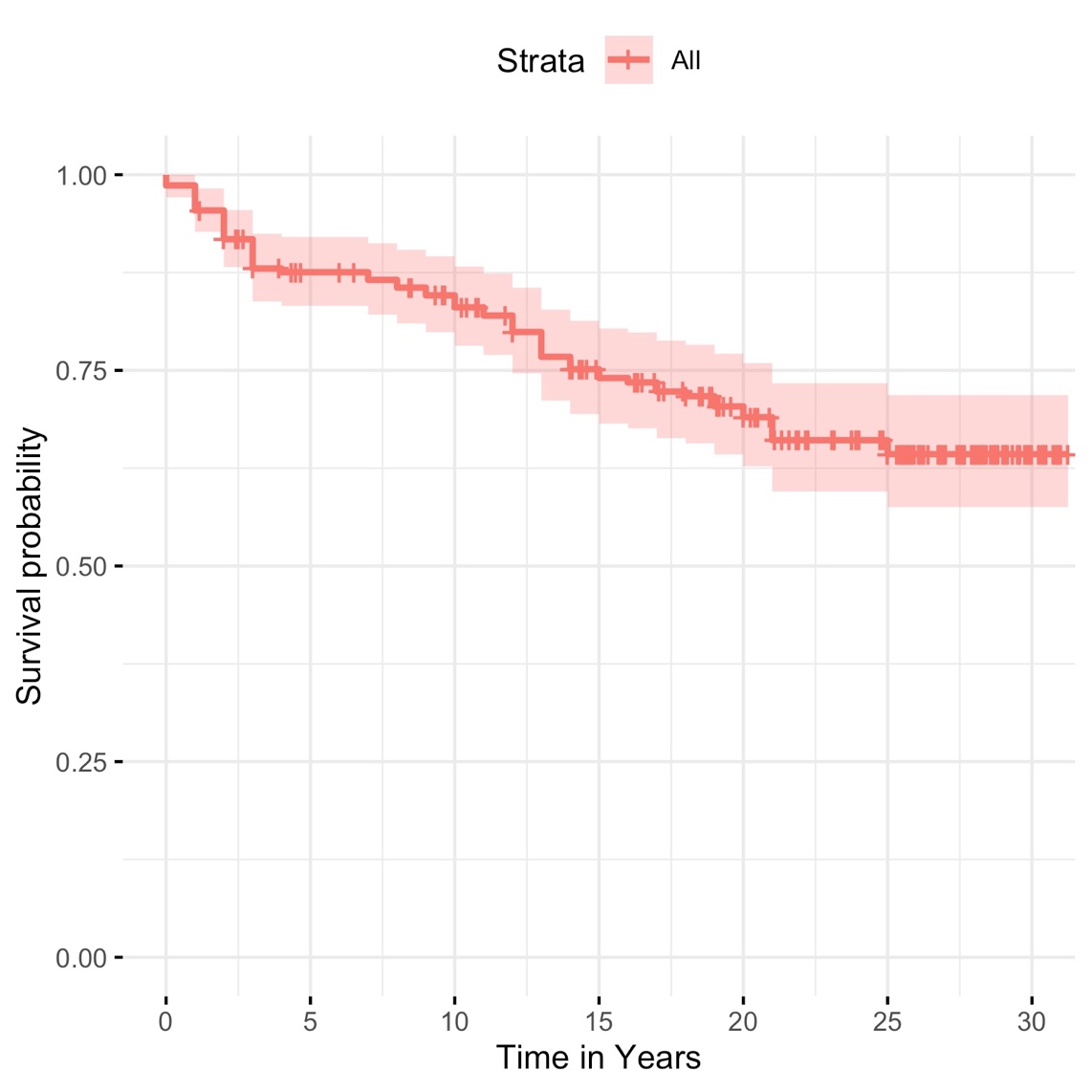

**Appendix B**

Individual Cox Regression Models

Cox Regression- Child Characteristics

| **Variable** | **Hazard Ratio** | **95% CI** | **P-value** |
| --- | --- | --- | --- |
| Gender | 1.23 | [0.62, 2,46] | 0.54 |
| Birth Order |  |  |  |
| Youngest Child | 0.36 | [0.19, 0.69] | 0.002 |
| Middle Child | 0.21 | [0.08, 0.50] | 0.0003 |
| Oldest Child | 0.30 | [0.13, 0.70] | 0.005 |
| ASD Diagnosis (ever) | 2.20 | [0.82, 5.93] | 0.12 |
| Verbal IQ | 1.00 | [0.99, 1.02] | 0.52 |
| CSS Social Affect | 1.07 | [0.96, 1.20] | 0.24 |
| CSS RRB | 0.94 | [0.84, 1.05] | 0.26 |
| Vineland Daily Living | 0.99 | [0.97, 1.01] | 0.38 |

*Note. CSS Calibrated Severity Score; reference group for Birth Order is Only Child; reference group for Gender is Male.

Cox Regression- Parent Characteristics

| **Variable** | **Hazard Ratio** | **95% CI** | **P-value** |
| --- | --- | --- | --- |
| Race | 2.11 | [1.24, 3.62] | 0.006 |
| Mother’s Education | 2.37 | [1.34, 4.18] | 0.003 |
| Race*Education | 10.47 | [1.27, 86.28] | 0.03 |
| Number of Siblings | 0.86 | [0.67, 1.10] | 0.23 |
| Mother’s Age | 0.95 | [0.91, 0.99] | 0.03 |

*Note. Reference group for Race is White, reference group for Parent Education is less than college education.

**Appendix C**

Descriptive Information across Child’s Age at Divorce

|  | **Number of siblings, M (SD)** | **ADOS CSS, M (SD)** | **ADOS CSS SA, M (SD)** | **ASDOS CSS RRB, M (SD)** | **Best VIQ, M (SD)** | **Best NVIQ, M (SD)** | **VABS Daily Living, M (SD)** |
| --- | --- | --- | --- | --- | --- | --- | --- |
| Divorce between 0-5 (n=27) | 1.5 (1.17) | 6.4 (2.77) | 6.72 (2.78) | 6.16 (2.84) | 59.37 (34.96) | 64.52 (29.84) | 63.92 (14.37) |
| ND between 0-5 (n=179) | 1.90 (1.29) | 5.94 (2.98) | 5.98 (2.91) | 6.56 (2.75) | 60.16 (38.19) | 67.52 (33.41) | 60.09 (16.38) |
| Divorce between 5-10 (n=9) | 2.22 (1.3) | 6.89 (3.1) | 6.78 (3.03) | 7.33 (2.96) | 43.56 (31.15) | 49.78 (27.84) | 54.00 (10.25) |
| ND between 5-10 (n= 169) | 1.89 (1.31) | 5.86 (2.97) | 5.91 (2.90) | 6.49 (2.73) | 61.40 (38.30) | 68.77 (33.51) | 60.85 (16.54) |
| Divorce between 10-15 (n=17) | 2.53 (1.8) | 7.41 (2.0) | 7.12 (1.9) | 7.65 (2.74) | 47.35 (36.03) | 57.47 (34.64) | 54.29 (14.95) |
| ND between 10-15 (n= 141) | 1.85 (1.23) | 5.55 (3.3) | 5.64 (2.98) | 6.25 (2.73) | 64.92 (37.68) | 72.77 (32.45) | 62.28 (16.29) |
| Divorce between 15-30 (n=14) | 1.79 (0.8) | 5.64(3.15) | 5.93 (3.32) | 6.14 (2.63) | 73.86 (33.26) | 87.46 (29.58) | 68.58 (15.49) |
| ND between 15-30 (n= 72) | 2.01 (1.36) | 6.13(2.89) | 6.10 (2.82) | 6.68 (2.66) | 59.07 (37.11) | 68.36 (32.43) | 61.40 (16.42) |

Note. ND= no divorce, ADOS= Autism Diagnostic Observation Schedule, CSS= Calibrated Severity Score, SA= Social Affect, RRB= Restricted and Repetitive Behavior
